# Neonatal amygdala microstructure and structural connectivity are associated with autistic traits at 2 years of age

**DOI:** 10.1101/2024.11.29.24318196

**Authors:** Kadi Vaher, Samuel R Neal, Manuel Blesa Cábez, Lorena Jiménez-Sánchez, Amy Corrigan, David Q Stoye, Helen L Turner, Rebekah Smikle, Hilary Cruickshank, Magda Rudnicka, Mark E Bastin, Michael J Thrippleton, Rebecca M Reynolds, James P Boardman

## Abstract

**Background:** Prenatal exposure to maternal stress is linked to behavioural and neurodevelopmental disorders in childhood. Maternal hair cortisol concentration in pregnancy associates with neonatal amygdala microstructure and structural connectivity ascertained from MRI, suggesting that amygdala development is sensitive to the impact of antenatal stress via hypothalamic-pituitary-adrenal axis. Here, we investigate whether amygdala microstructure and/or connectivity associate with neurodevelopment at 2 years of age.

**Methods:** 174 participants (105 very preterm) underwent brain MRI at term-equivalent age and assessment of neurodevelopment, autistic traits, temperament, and executive function at 2 years corrected age. We calculated diffusion tensor imaging and neurite orientation dispersion and density imaging metrics for left and right amygdalae. Structural connectivity was measured by mean fractional anisotropy from the amygdalae to 6 ipsilateral regions of interest (insula, putamen, thalamus, inferior temporal gyrus, medial orbitofrontal cortex, rostral anterior cingulate cortex). We used linear regression to model amygdala-outcome associations, adjusting for gestational age at birth and at scan, sex, maternal education, and maternal postnatal depression score. Network-based statistics (NBS) was used for a whole-brain analysis.

**Results:** After adjusting for multiple comparisons, lower amygdala mean diffusivity bilaterally (left: β=-0.32, p=0.026, right: β=-0.38, p=0.012), higher left amygdala neurite density index (β=0.35, p=0.026), and increased left amygdala-putamen connectivity (β=0.31, p=0.026) associated with higher autistic traits. NBS revealed amygdala-involving networks associated with cognition and surgency temperament trait among preterm infants. Other neurodevelopmental outcomes did not significantly associate with amygdala imaging features.

**Conclusions:** Microstructural variation in the neonatal amygdala may be important in the development of autistic traits.

## INTRODUCTION

Exposure to prenatal stress, commonly experienced by women during pregnancy (1), associates with cognitive, behavioural and mental health conditions across the life course, including higher rates of anxiety and depression, negative affectivity, autistic traits, and diagnoses such as attention deficit hyperactivity disorder and schizophrenia (2,3), often in a sexually dimorphic manner (4). Dysregulation of the hypothalamic-pituitary-adrenal (HPA) axis may be one major mechanism whereby maternal stress influences offspring brain development (5). The amygdala is central to emotional and social information processing (6), and is a compelling target for neurodevelopmental programming via the HPA axis. The amygdala develops early in gestation (7), has a high concentration of glucocorticoid receptors (8), and undergoes protracted development into adolescence (9,10).

In offspring exposed to prenatal stress, magnetic resonance imaging (MRI) has revealed altered structural and functional connectivity in brain networks involving the amygdala (11,12). Specifically, we previously showed that maternal hair cortisol concentration sampled soon after birth (a marker of cortisol levels over the last 3 months of pregnancy) associates with neonatal amygdala microstructure and structural connectivity in a sexually dimorphic manner (13). These findings reinforce previous associations between maternal salivary cortisol levels during pregnancy and functional connectivity of amygdala-involving networks in neonates (14) and amygdala volumes in childhood (15). Furthermore, self-reported/perceived maternal prenatal stress correlates with amygdala volume (16), grey matter microstructure (17), and structural (18) and functional connectivity (18–20).

Neuroimaging studies have demonstrated associations between amygdala anatomy and numerous neurodevelopmental and psychiatric conditions such as autism spectrum disorder, anxiety and schizophrenia (21). Methodological advances in MRI techniques over the past decade have further enabled characterisation of the neonatal amygdala in relation to childhood outcomes. Increased amygdala volume at term-equivalent age has been associated with poorer working memory (22) and heightened fear response in toddlers (23). Furthermore, altered resting-state functional connectivity of the neonatal amygdala to other brain regions correlates with fear and sadness (24,25), increased internalising behaviour (14,26), atypical social communication (27), and socio-emotional development in childhood (28). Some amygdala-behaviour associations have been shown to be sexually dimorphic (14,22,29), though evidence remains mixed. Most of these studies have been conducted using relatively small sample sizes (median n=58) and few have included infants born very preterm. This is a limitation because stress-amygdala interactions are expected to operate across the whole of gestation, and because people born preterm are disproportionally affected by cognitive and socio-emotional difficulties and have a higher prevalence of neurodevelopmental and psychiatric diagnoses (30,31). Importantly, current literature leaves uncertainty about the associations between neonatal amygdala microstructure or structural connectivity – features we previously demonstrated to be associated with prenatal maternal cortisol levels (13) – and subsequent neurodevelopment. To address these knowledge gaps, we investigated relationships between neonatal amygdala microstructure and structural connectivity, and a range of neurodevelopmental outcomes at two years of age in a sample enriched for prematurity.

## METHODS AND MATERIALS

### Participants

Participants were very preterm (gestational age (GA) at birth ≤32 completed weeks) and term-born infants recruited to the prospective longitudinal Theirworld Edinburgh Birth Cohort study (32). Infants were born and recruited at the Royal Infirmary of Edinburgh, UK, between September 2016 and September 2021. The study was conducted according to the principles of the Declaration of Helsinki, and ethical approval was obtained from the UK National Research Ethics Service (South East Scotland Research Ethic Committee 16/SS/0154). Parents provided written informed consent.

Exclusion criteria were congenital malformation, chromosomal abnormality, cystic periventricular leukomalacia, haemorrhagic parenchymal infarction, and post-haemorrhagic ventricular dilatation. These criteria mean the cohort is representative of most survivors of modern intensive care practices (32). The conventional neonatal MRI results from the cohort have been reported previously (33).

Here, we included participants who had data available for multimodal brain MRI acquired at term-equivalent age and at least one outcome measure at 2 years of corrected age.

### Demographic and clinical information

Participant demographic and clinical information was collected through questionnaires and medical records. Please see Table S1 for details.

### Magnetic resonance imaging

#### Image acquisition

Infants were scanned at term-equivalent age in natural sleep at the Edinburgh Imaging Facility, Royal Infirmary of Edinburgh, University of Edinburgh, UK, using a Siemens MAGNETOM Prisma 3T MRI clinical scanner (Siemens Healthcare Erlangen, Germany) as previously described (32,34,35). See Supplementary information for details.

#### Image pre-processing and feature extraction

Image data were processed and networks constructed as previously described (13,34,35), see details in the Supplementary information.

##### Microstructural features

Diffusion tensor imaging (DTI) and Neurite orientation dispersion and density imaging (NODDI) maps were calculated in the dMRI processed images to obtain fractional anisotropy (FA), mean diffusivity (MD), neurite density index (NDI) and orientation dispersion index (ODI). The DTI model was fitted in each voxel using the weighted least-squares method DTIFIT as implemented in FSL (36) using only the b = 750 s/mm^2^ shell. NODDI metrics were calculated using all shells and the recommended values for neonatal grey matter of the parallel intrinsic diffusivity (1.25 μm^2^/ms) (37,38). The mean FA, MD, ODI, and NDI were calculated for the left and right amygdalae from the Melbourne Children’s Regional Infant Brain atlas parcellation (39).

##### Structural connectivity features

We selected four unilateral connections of interest from the amygdalae as predictors of neurodevelopmental outcomes due to their significant associations with either the interaction effect between maternal hair cortisol concentration and infant sex, or with maternal hair cortisol concentration in sex-stratified analyses in our previous work (13): putamen, thalamus, insula, and inferior temporal gyrus. We selected two additional connections from the amygdalae to the medial orbitofrontal cortex and rostral anterior cingulate cortex due to their associations with behavioural outcomes in other paediatric studies (24,25,29,40). Mean FA for each connection was calculated as an indicator of structural connectivity strength/integrity. Segmentation of the amygdalae and the 6 connected regions of interest are visualised in Figure 1.

**Figure 1.**
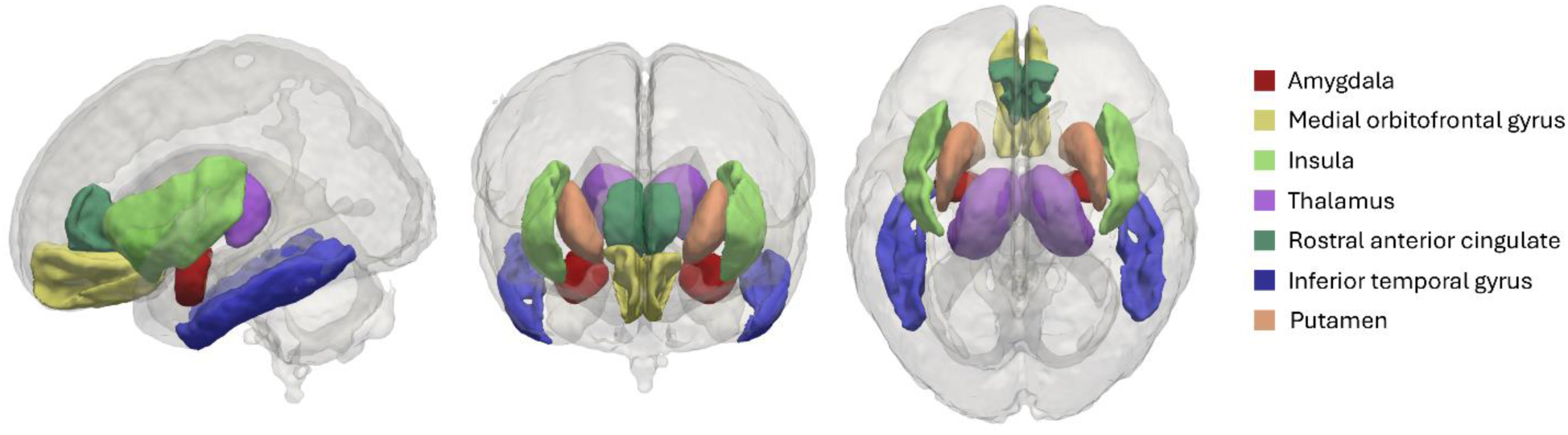
Segmentations of the amygdalae and connected regions of interest in the ENA50 neonatal atlas space. Shown in lateral (left), anterior (centre) and superior (right) views.

#### Follow-up appointment and behavioural testing

At 2 years of corrected age, we collected information about participants’ neurodevelopment. Primary caregivers were asked to complete the following questionnaires to assess neurodevelopment across domains.

##### Early Childhood Behavioural Questionnaire (ECBQ

ECBQ assesses temperament between the ages of 18 and 36 months (41). From the questionnaire data, we calculated the three established factors that represent broad dimensions of temperament: Negative Affectivity, Surgency/Extraversion, and Effortful Control. Higher scores indicate higher prevalence of behaviours indicative of these traits.

##### Behaviour Rating Inventory of Executive Function, Preschool Version (BRIEF-P)

BRIEF-P assesses the range of behavioural manifestations of infant’s executive function (42). We calculated the Global Executive Functioning, with higher scores indicative of poorer executive functioning. Infants with elevated negativity or high inconsistency ratings (n=4) were excluded from the analyses.

##### Quantitative Checklist for Autism in Toddlers (Q-CHAT)

Q-CHAT assesses frequency of behaviours also observed in autism spectrum conditions in toddlers aged 18 to 30 months with the aim of identifying those who should be referred for diagnostic assessment (43). We obtained a total Q-CHAT score, with higher scores indicative of higher frequency of autistic traits. Missing questionnaire items were replaced with zeros to calculate the total Q-CHAT score, assuming at least one item was completed (44). Data were imputed for 12 infants with a range of 1-3 questionnaire item values missing.

##### Bayley Scales of Infant and Toddler Development, Third Edition (Bayley-III)

Preterm infants additionally completed the Bayley-III (45) with a trained clinician at 2 years of corrected age (45) as part of their routine National Health Service developmental follow-up appointment. From medical records, we obtained the standardised composite scores for four domains measured with Bayley-III: cognitive, language, adaptive, and social-emotional development, with higher scores indicative of better functioning in these domains. To reduce the number of tests, the motor domain was omitted as amygdala is not a dominant component of the motor system.

### Statistical analysis

Statistical analyses were performed in R (version 4.3.2) (46).

#### Covariates

The following covariates were included based on known associations with neonatal brain structure and/or neurodevelopment and their correlation with at least one outcome measure in the current sample: GA at birth, sex, ethnicity, maternal postnatal depression score (dichotomised as ≤10), maternal age and maternal education. We also adjusted for GA at scan due to its strong correlation with brain MRI metrics. See further details on covariate selection in Supplementary information.

In the interest of constructing a parsimonious model, we removed infant ethnicity and maternal age because these variables associated with only one outcome. Full and sparser model comparison using analysis of variance (ANOVA), and Akaike and Bayesian information criteria revealed that the sparser model fit the data as well (not shown). Therefore, the final covariates included in the analysis were: GA at birth, infant sex, GA at scan, maternal final education qualification, and postnatal depression score.

#### Main analyses

Separate multiple linear regression models were constructed to test for associations between each neurodevelopmental outcome (dependent variable) and MRI feature (independent variable) for the left and right amygdala. In total, we tested 9 outcome scores and 10 MRI features for each hemisphere, resulting in 180 models.

To maximise sample size, analyses were performed on the subset of infants with available data for each outcome, resulting in nine subsets of our study sample.

Given prior evidence, we originally considered infant sex*MRI feature interaction term in the models. The interaction term was not consistently associated with the outcomes (p < 0.05 in 3/180 models) and model comparisons did not indicate improvement in fit when including the interaction term, except for the 3 models. Therefore, models without the interaction term are presented in the Results.

We tested model assumptions of linearity, homoscedasticity, and normality of residuals by visually inspecting the model diagnostic plots; presence of multicollinearity was determined by calculating the variance inflation factor for each covariate.

All continuous variables were z-transformed (scaled) prior to fitting the models. We report standardised β coefficients and 95% confidence intervals (CI). We controlled the false discovery rate at 0.05 using the Benjamini-Hochberg (47) procedure applied separately to each outcome measure.

Given that Bayley-III data were only available for preterm infants, we performed subgroup analyses separately within the term and preterm groups following the same model specifications to investigate gestation-dependent associations.

#### Sensitivity analyses

Three sensitivity analyses were carried out.

First, because the variation in amygdala microstructure and its connectivity to other brain regions may be transmitting maternal factors to infant outcomes, we repeated all main significant models excluding maternal factors (education, postnatal depression score). The full model and the one without maternal factors were compared using ANOVA, Akaike and Bayes information criteria, the adjusted R^2^, and the standardised β coefficients of the amygdala features.

Second, this dataset contained 14 twin pairs (13 preterm). We re-ran models with significant amygdala-outcome associations, including one randomly selected twin from each pair.

Third, to explore the robustness of the amygdala-outcome associations, we repeated the models with significant amygdala-outcome associations in the sample of our previous work showing associations between maternal hair cortisol and amygdala microstructure/connectivity (13). Out of the n=78 in the previous study, 39 (8 preterm [20.5%]) had data available for Q-CHAT.

#### Network based statistics

We performed network-based statistics (NBS) (48) to identify whether the amygdala might be part of a larger network that is associated with outcomes. Edge-wise comparison of connectivity matrices was performed using *connectomestats* within MRtrix3 (49). NBS results are highly dependent on the primary test-statistics threshold. Therefore, given the lack of an established test-statistic threshold to use in NBS analysis of neonatal/paediatric data, we tested a range of values (t = 1.5–3.5, with 0.1 increments; i.e. 21 experiments) (50). All other parameters were set to their default values. The NBS analyses, similarly to main regression models, were adjusted for GA at birth, infant sex, GA at scan, maternal final education qualification, and risk for postnatal depression; continuous variables were z-transformed.

We took a multi-verse approach, reasoning that confidence in the implicated connections is increased if they appear under several test-statistic values. Therefore, focussing on results for which a connection with amygdala was part of the statistically significant outcome-associated network (p < 0.05), for each connection we calculated the proportion of experiments where it was part of the associated network. Then, we filtered this to focus on and visualise only those connections that appeared in at least 7/21 (33.3%) of experiments, considering them as consistent and less dependent on the selected statistic threshold. Similarly to regression models, we ran subgroup analyses in term and preterm groups separately.

### Data and code availability

Data used for analysis in this work are deposited in Edinburgh DataVault (51) (https://doi.org/10.7488/e65499db-2263-4d3c-9335-55ae6d49af2b). Requests for access for these as well as raw neuroimaging data will be considered under the study’s Data Access and Collaboration policy and governance process (https://www.ed.ac.uk/centre-reproductive-health/tebc/about-tebc/for-researchers/data-access-collaboration).Code used for the data analysis in this paper is available on GitLab (https://git.ecdf.ed.ac.uk/jbrl/amy-neurodev).

## RESULTS

### Participant characteristics

174 participants (characteristics shown in Table 1) had available data for multimodal neonatal brain MRI and at least one outcome measure collected at 2 years of life. The distributions and intercorrelations of the 2-year outcome variables are presented in Figure 2. Q-CHAT scores were higher in preterm compared to term infants (t =-4.345, p < 0.001) and ECBQ negative affectivity trait was slightly lower in the term group (Figure S1).

**Figure 2.**
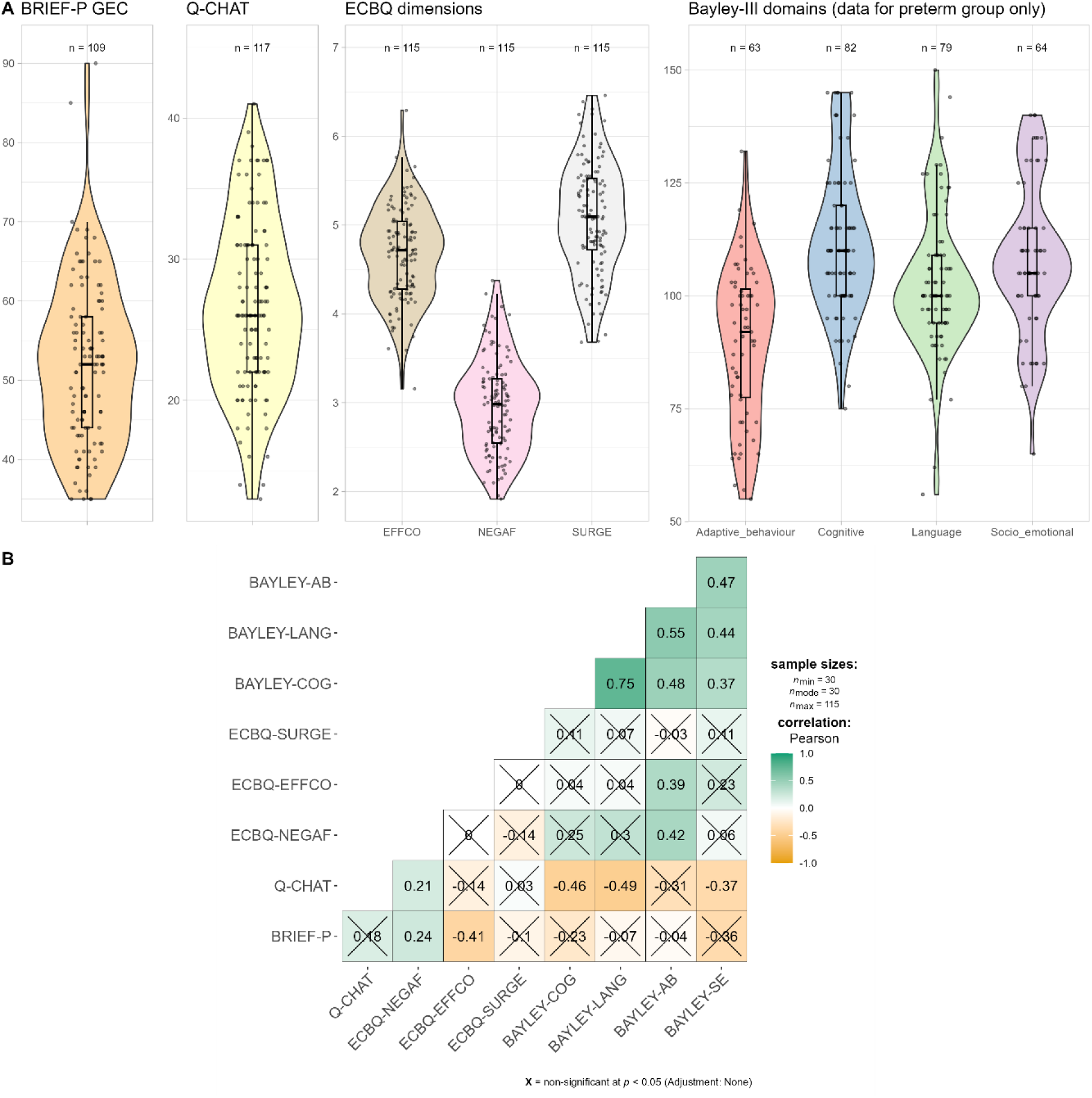
Distributions (A) and Pearson correlations (B) of 2-year outcome variables. Crossed out boxes indicate statistically non-significant correlations. BRIEF-P = Behavior Rating Inventory of Executive Function, Preschool, GEC = global executive composite, Q-CHAT = Quantitative Checklist for Autism in Toddlers; ECBQ = Early Childhood Behavior Questionnaire; EFFCO = effortful control, NEGAF = negative affectivity, SURGE = surgency, AB = adaptive behaviour, LANG = language, COG = cognitive, SE = socio-emotional.

**Table 1.** Summary of infant and maternal demographics.

| Characteristic | $n = 174^*$ |
| --- | --- |
| **Infant** |  |
| Male | 103 (59%) |
| Gestation at birth (weeks) | 31.43 [22.14 to 42.14] |
| Preterm | 105 (60%) |
| Birthweight (g) | 1690 [370 to 4560] |
| Birthweight z-score | 0.20 (1.03) |
| Gestation at MRI (weeks) | 41.20 (1.53) |
| **Maternal** |  |
| Age at booking (years) | 33.4 (5.0) |
| BMI at booking ($\text{kg}/\text{m}^2$) | 25.1 [16.4 to 47.0] |
| University or postgraduate education | 121 (70%) |
| Edinburgh Postnatal Depression Scale $\geq 10$ | 42 (24%) |

\*Data are presented as $n$ (%) for categorical variables, mean (SD) for continuous and normally distributed variables, or median [range] for continuous non-normally distributed variables.

### Amygdala microstructure and structural connectivity associate with autistic traits

After adjustment for multiple tests, amygdala microstructural indices showed statistically significant associations only with autistic traits (Figure 3A): MD in the amygdala bilaterally was negatively (left: β =-0.316, 95% CI [-0.535 to-0.097]; right: β =-0.375, 95% CI [-0.586 to-0.165]) and NDI in the left amygdala positively (β = 0.346, 95% CI [0.113 to 0.579]) associated with Q-CHAT scores. The structural connectivity (mean FA) between left amygdala and putamen positively associated with Q-CHAT scores (β = 0.311, 95% CI [0.102 to 0.521]; Figure 3B). We found no statistically significant associations between amygdala features and temperament (ECBQ), executive function (BRIEF-P) or Bayley-III composite scores (Table S2).

**Figure 3.**
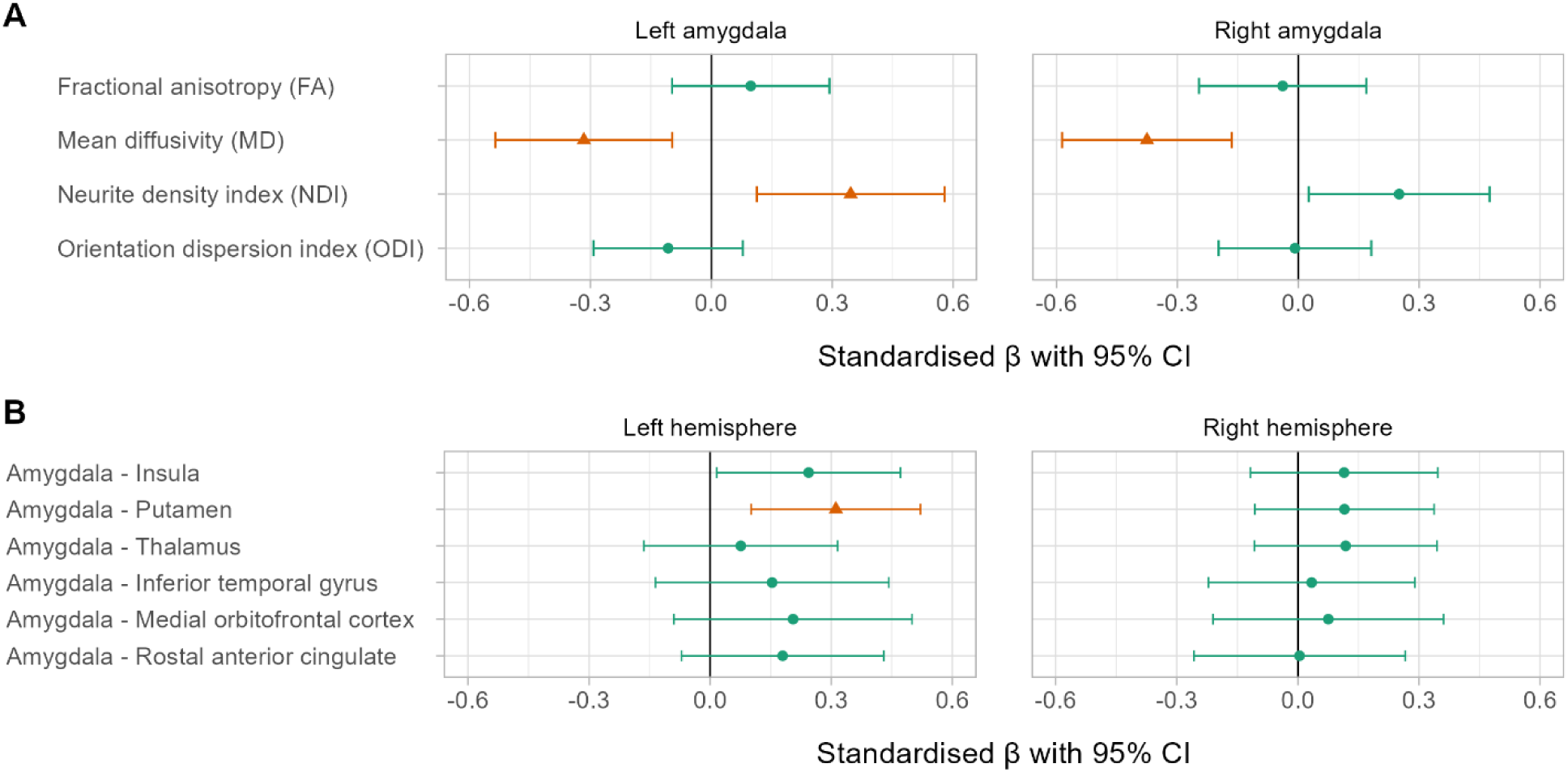
Amygdala microstructure (A) and structural connectivity (B) associations with Q-CHAT scores. Models are adjusted for gestational age at birth and at scan, infant sex, maternal education (university/postgraduate degree vs lower) and high postnatal depression screening score (Edinburgh Postnatal Depression Scale ≥ 10). Coefficients shown as orange lines and triangular shapes indicate statistically significant associations after adjustment for multiple comparisons using the Benjamini-Hochberg procedure.

### Sensitivity analyses

The association strength between amygdala microstructure/connectivity and outcomes was minimally altered when excluding maternal factors from the model (average change in standardised β of the imaging predictor: 0.018; **Error! Reference source not found.**), indicating that amygdala MRI features are associated with autistic traits independent of maternal education and postnatal depression score.

The four amygdala microstructure/connectivity and Q-CHAT associations remained statistically significant when excluding one twin randomly from the sample (n=108 with Q-CHAT data), with little change to the regression coefficients.

We repeated the significant amygdala-outcome models in a smaller subset of participants overlapping with our previous publication (13) who had data available for cortisol, MRI and Q-CHAT. The standardised β-s were similar and within the 95% confidence intervals of the larger sample (Figure 4). This consistency suggests that the observed amygdala-outcome associations are robust and not dependent on sample size, providing additional confidence in the reliability of these findings.

**Figure 4.**
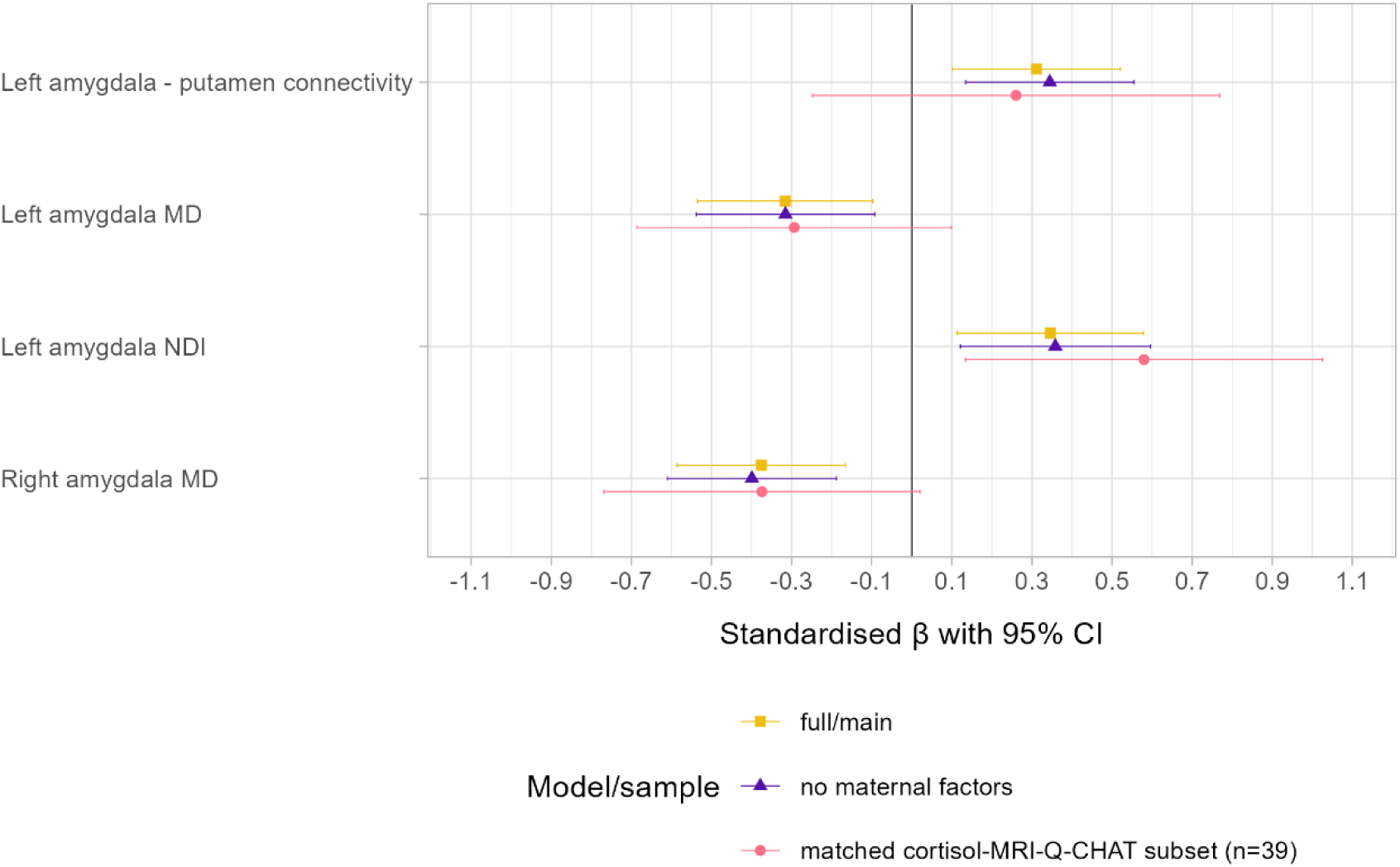
Amygdala microstructure and structural connectivity associations with Q-CHAT in sensitivity analyses in comparison with the main model. Full/main indicates the original model associating amygdala features with Q-CHAT adjusting for gestational age at birth, infant sex, gestational age at scan, maternal final education qualification, and risk for postnatal depression; no maternal factors indicates the original model but excluding maternal final education qualification and risk for postnatal depression; matched cortisol-MRI-Q-CHAT subset indicates the original model in a smaller subset who had data available for MRI, Q-CHAT, and prenatal maternal cortisol.

### Subgroup analysis

The Q-CHAT negative associations with amygdala MD were similar in term and preterm group (Table S3). In contrast, the positive associations with NDI and left amygdala-putamen connectivity were stronger in the term group. In the term group, 2-year Q-CHAT scores were further correlated with left amygdala ODI as well as positively with some other amygdala connections.

### Network-based statistics

Given the largely null fundings in our main analyses for amygdala structural connectivity, we used NBS to determine if amygdala is part of larger brain networks correlated with outcomes. We found five connections from the right amygdala and one connection from the left amygdala that were part of a brain-wide network which positively correlated with Bayley-III cognitive scores among preterm infants (Figure 5). These six connections were part of the associated network at test-statistic thresholds from 1.5 to 2.1. Amygdala connections were not part of consistently associated networks for other 2-year outcome measures across the entire sample.

**Figure 5.**
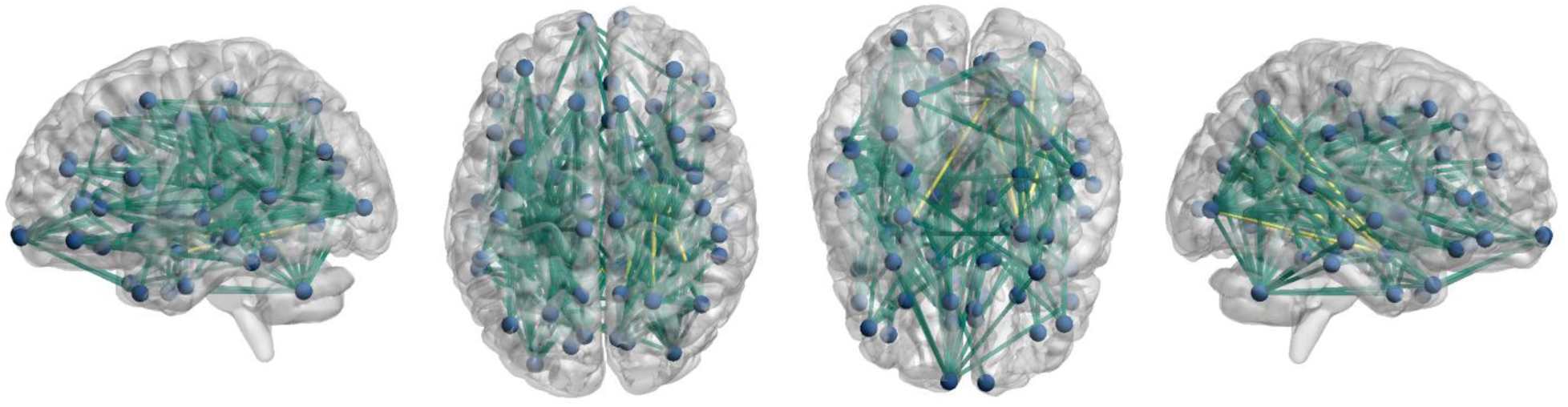
Illustration of network-based statistics results for Bayley-III cognitive domain. Connections that appear as significant in at least 33.3% of experiments are shown; amygdala connections (right amygdala to right cuneus, right superior parietal cortex, right inferior parietal cortex, right lateral occipital cortex and right precuneus, and left amygdala to right lingual cortex) are shown in yellow.

With regards to Q-CHAT, among the term group, there were 3 connections from the right and 8 connections from the left amygdala that were consistently, mostly at test-statistic thresholds from 1.5 to 2.3, part of a network whose connectivity positively correlated with Q-CHAT scores (Figure 6A). In contrast, among the preterm group, NBS identified that amygdala was part of a network whose connectivity negatively correlated with Q-CHAT scores. The consistent connections, mostly at test-statistic thresholds from 2.1 to 2.9, included 4 connections from the left and right amygdalae (Figure 6B).

**Figure 6.**
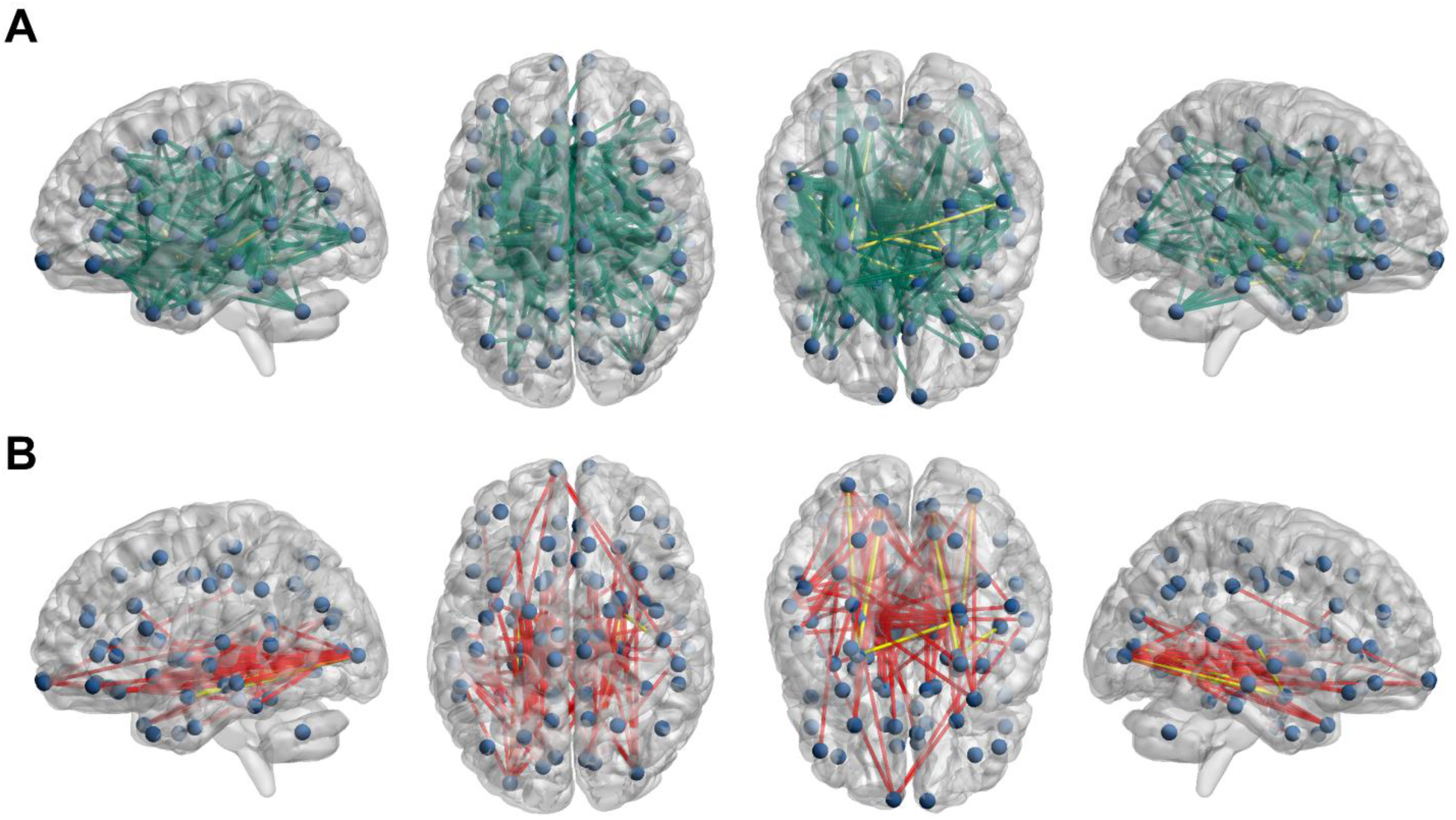
Illustration of network-based statistics results for Q-CHAT in term (A) and preterm (B) groups. Connections that appear as significant in at least 33.3% of experiments are shown; amygdala connections are shown in yellow. In the term group (A), connections were positively, while in the preterm group (B), connections were negatively correlated with Q-CHAT scores. In the term groups the amygdala connections are: from the right amygdala to left caudal anterior cingulate, left isthmus cingulate and left transverse temporal gyrus, and from the left amygdala to left banks of the superior temporal sulcus, left lingual cortex, left insula, left caudate, left putamen, right caudate, right inferior temporal gyrus and right supramarginal gyrus. In the preterm group, the amygdala connections are: from the left amygdala to left lateral occipital cortex, left lingual cortex, left pericalcarine cortex and the right hippocampus, and from the right amygdala to the right lateral occipital cortex, right lingual cortex, right pericalcarine cortex and right transverse temporal cortex.

Subgroup analyses revealed that the amygdala is part of a network whose connectivity is positively correlated with ECBQ surgency dimension in preterm infants (Figure S2). This includes both inter-and intra-hemispheric connections.

### Maternal hair cortisol and Q-CHAT

Finally, we explored to what extent maternal hair cortisol concentration correlates with Q-CHAT scores. There was no correlation between maternal hair cortisol concentration and Q-CHAT score across the entire sample (Figure S3A). However, there may be sex-specific relationships between maternal cortisol and Q-CHAT such that there is a negative correlation in male and positive correlation in female infants (Figure S3B; β_interaction_ =-0.661, p = 0.043, adjusted for GA at birth, n=39).

## DISCUSSION

By combining multimodal brain MRI and developmental outcome data, our study shows that neonatal amygdala microstructure and structural connectivity to putamen – previously linked to maternal cortisol levels in pregnancy – associate with autistic traits at 2 years of age. Whole-brain analyses revealed an amygdala-involving network associated with cognitive scores in preterm infants and suggestions for gestation-dependent associations between brain connectivity and autistic traits.

Previous work has shown amygdala enlargement in autistic children, with positive correlations between amygdala volume and autistic traits (21,52–54), followed by reduced growth later, resulting in amygdala that is smaller than or of equal size to typically developing individuals in adolescence and adulthood (9,21). Histologic analyses have further found initial excess of amygdala neurons/neurites in autistic children (55). Our findings of higher NDI and lower MD in association with higher autistic traits appear consistent with this. We also found evidence that stronger neonatal amygdala-putamen connectivity in the left hemisphere may be positively correlated with 2-year autistic traits. Differences in anatomy and functional connectivity of basal ganglia and the striatum, including putamen, have been shown to associate with autistic behaviours, diagnosis, or genetic risk scores (56–58).

Our term/preterm segregated analyses, in both targeted regression as well as whole-brain NBS analyses, suggest that there are gestation-dependent associations between amygdala features and autistic traits. Specifically, higher structural connectivity of the neonatal amygdala in term infants was associated with a higher frequency of autistic traits in toddlerhood, but in the preterm group it was associated with a lower frequency. Varying relationships in term and preterm infants are plausible given that the multitude of adverse co-exposures of preterm birth can further impact microstructural properties of the neonatal brain (59), including amygdala and its connections. The amygdala-insula connectivity correlation with autistic traits in the term group is interesting in the context of previous work showing that lower functional connectivity between left amygdala and insula in neonates is correlated with family history of autism as well as higher social risk scores at 1 year of age (27).

To link our previous findings of sexually dimorphic cortisol-amygdala microstructure associations (13) with the main findings of the current study, in a smaller overlapping sample we found evidence for a sex-specific relationship between maternal hair cortisol levels and Q-CHAT scores such that there was a negative correlation in males and a positive correlation in females. This is in line with previous studies suggesting a sex interaction effect between maternal blood cortisol levels in pregnancy and autistic behaviours in childhood, with particularly prominent associations in males, though there is mixed evidence about the direction of this association (60,61). However, direct comparisons of the three studies are complicated given their methodological differences in assessing autistic behaviours and cortisol. Our analyses did not find evidence for a sex-by-amygdala interaction, suggesting that the extent to which the cortisol-associated neonatal amygdala MRI features correlate with subsequent autistic traits is similar in males and females. These results together raise the possibility of a sex-moderated mediation effect such that higher prenatal cortisol levels in males may lead to changes in amygdala that are subsequently linked with lower frequency of autistic traits, whilst in females higher prenatal cortisol associates with changes in amygdala which in turn relate to higher frequency of autistic traits. This is akin to a report by Graham et al. (14) who found that neonatal amygdala connectivity was similarly correlated with internalising behaviours in male and female infants, but that amygdala connectivity mediated maternal cortisol-behaviour associations in females only. This interpretation should be approached with caution because the analyses discussed here stem from only partially overlapping samples. Therefore, future larger studies are required to formally test this hypothesis of a sex-moderated mediation effect of the neonatal amygdala for autistic traits.

Whole brain connectivity analyses revealed increased neonatal structural connectivity of brain networks in a positive association with surgency trait and Bayley-III cognitive scores at 2 years of age in preterm infants. Amygdala was part of these networks, but not the sole contributor, and the 6 cortisol-associated connections of interest were not included in these networks. Increased amygdala functional connectivity in infancy has been mainly associated with higher negative affectivity traits such as fear and sadness (24,25), internalising behaviours (14,26), and lower positive affectivity (62). In contrast, in a previous study, neonatal amygdala structure or whole brain structural connectivity did not associate with attachment behaviours at 9 months of age (63), though it is salient that the correlations between temperament traits and attachment are weak (64). Therefore, our results appear in contrast with previous functional findings; yet, it is uncertain to what extent functional and structural connectivity measures correlate in neonates as some studies show no correlations in networks involving the amygdala (28) and there is spatially varying structural-functional connectivity coupling across the brain (65). The Bayley-III results are in line with previous studies showing positive correlations between white matter FA or structural connectivity across the brain and cognition following preterm birth (e.g. (66,67)). Future studies should determine if similar relationships are present in the term group.

The study has some limitations and several avenues for future research can be considered. First, our cohort is enriched for very preterm infants who are at risk of atypical development in broad domains that can include autistic traits, indicating that further work is required to understand whether findings generalise. Preterm infants are more likely to be diagnosed with autism (68) and because our population is enriched for preterm birth, higher prevalence of autistic traits in the study group could be expected. Accordingly, we found higher Q-CHAT scores in preterm compared to the term group in our cohort. However, only two preterm participants had scores >39, a screening cut-off point shown to maximise sensitivity and specificity for autism diagnosis (43), and the Q-CHAT scores in our sample (mean = 27, range 13 to 41) appear lower than those recently reported in the term-dominant dHCP cohort (mean = 30.5, range 8 to 70) (69). Q-CHAT instrument has also been shown to have poor positive predictive value for autism, thus, the scores may instead reflect developmental/behavioural differences (43,69). Therefore, the extent to which these findings generalise to the prediction of future autism diagnosis remains to be tested in future studies.

Second, the mothers of 70% of our participants had university/postgraduate degrees, so the dataset is skewed towards higher socioeconomic status, which can impact early child development. Maternal education was included as a covariate, but further studies are needed to confirm if the findings generalise to the wider population. Third, our study primarily used parent-completed questionnaires for outcome measures, which, despite being at risk for reporting biases, are considered to have good ecological validity (70). Future work should contextualise these results with complementary direct measures including diagnostic assessment tools.

In summary, our results suggest that variation in the microstructure of the neonatal amygdala may predict the frequency of autistic traits in infants born across the gestational range, which may be relevant for the emergence of neurodevelopmental conditions that include autistic traits in their phenomenology.

## Data Availability

Data used for analysis in this work are deposited in Edinburgh DataVault (https://doi.org/10.7488/e65499db-2263-4d3c-9335-55ae6d49af2b). Requests for access for these as well as raw neuroimaging data will be considered under the study's Data Access and Collaboration policy and governance process (https://www.ed.ac.uk/centre-reproductive-health/tebc/about-tebc/for-researchers/data-access-collaboration). Code used for the data analysis in this paper is available on GitLab (https://git.ecdf.ed.ac.uk/jbrl/amy-neurodev).

## ACKNOWLEDGEMENTS

For the purpose of open access, the author has applied a CC BY public copyright license to any Author Accepted Manuscript version arising from this submission. This work was supported by Theirworld (www.theirworld.org) and the Medical Research Council (MR/X003434/1). Participants were scanned in the University of Edinburgh Imaging Research MRI Facility at the Royal Infirmary of Edinburgh, which was established with funding from The Wellcome Trust, Dunhill Medical Trust, Edinburgh and Lothians Research Foundation, Theirworld, The Muir Maxwell Trust and other sources. KV and LJ-S were supported by the Translational Neuroscience programme, funded by Wellcome (Grant no. <u>108890/Z/15/Z</u>). RS is a student on the Translational Neuroscience PhD programme, funded by Wellcome (Grant no. 218493/Z/19/Z). HT is a PhD student in Clinical Brain Sciences and receives funding support from Simpson Special Care Babies charity. We are grateful to the families who consented to take part in the study and to the radiographers at the Edinburgh Imaging Facility at the Royal Infirmary of Edinburgh for infant scanning.

