## Supplementary information for "Neonatal amygdala microstructure and structural connectivity are associated with autistic traits at 2 years of age"

#### SUPPLEMENTARY METHODS

##### Image acquisition

Full imaging protocol is available in the study protocol paper (1). Briefly, a 16-channel phased-array paediatric head coil was used to acquire 3D T2-weighted SPACE images (T2w) (voxel size = 1 mm isotropic, TE = 409 ms and TR = 3200 ms; acquisition time = 2:13 min) and axial diffusion MRI (dMRI) data. dMRI images were acquired in two separate acquisitions to reduce the time needed to re-acquire any data lost to motion artifacts: the first acquisition consisted of 8 baseline volumes (b = 0 s/mm$^2$ [b0]) and 64 volumes with b = 750 s/mm$^2$; the second consisted of 8 b0, 3 volumes with b = 200 s/mm$^2$, 6 volumes with b = 500 s/mm$^2$ and 64 volumes with b = 2500 s/mm$^2$ (acquisition time = 4:29 + 5:01 min). An optimal angular coverage for the sampling scheme was applied (2). An acquisition of 3 b0 volumes with an inverse phase encoding direction was also performed (acquisition time = 0:28 min). All dMRI images were acquired using single-shot spin-echo echo planar imaging (EPI) with 2-fold simultaneous multislice and 2-fold in-plane parallel imaging acceleration and 2 mm isotropic voxels; all three diffusion acquisitions had the same parameters (TR/TE 3400/78.0 ms).

Infants were fed and wrapped and allowed to sleep naturally in the scanner. Pulse oximetry, electrocardiography and temperature were monitored. Flexible earplugs and neonatal earmuffs (MiniMuffs, Natus) were used for acoustic protection. All scans were supervised by a doctor or nurse trained in neonatal resuscitation. Each acquisition was inspected contemporaneously for motion artefact and repeated if there had been movement but the baby was still sleeping; dMRI acquisitions were repeated if signal loss was seen in 3 or more volumes.

##### Image pre-processing

dMRI processing was performed as follows: for each subject, the two dMRI acquisitions were first concatenated and then denoised using a Marchenko-Pastur-PCA-based algorithm (3); eddy current, head movement and EPI geometric distortions were corrected using outlier replacement and slice-to-volume registration (4–7); bias field inhomogeneity correction was performed by calculating the bias field of the mean b0 volume and applying the correction to all the volumes (8).

The T2w images were processed using the minimal processing pipeline of the developing human connectome project (dHCP) to obtain the bias field corrected T2w, the brain masks and the different tissue probability maps (9).

Finally, the mean b0 EPI volume of each subject was co-registered to their structural T2w volume using boundary-based registration (10).

Melbourne Children's Regional Infant Brain (M-CRIB) atlas parcellation (11) was used to obtain regional parcellation. The 10 manually labelled subjects of the atlas were registered to the bias field corrected T2w using rigid, affine and symmetric normalisation (SyN) (12). The registered labels of the 10 atlases were then merged using joint label fusion (13), resulting in a parcellation containing 84 regions of interest (ROIs).

##### Network construction and analysis

Tractography and fractional anisotropy-weighted connectomes were constructed as previously published (14). We performed anatomically constrained tractography using constrained spherical deconvolution (15,16). We used FA threshold of 0.1 to calculate the multi-tissue response function, followed by calculation of the average response functions. Then, multi-tissue fiber orientation distribution (FOD) was calculated (17) and global intensity normalisation on the FODs images was performed. Finally, the tractogram was created, generating 10 million streamlines, with a minimum length of 20 mm and a maximum of 200 mm and a cut-off of 0.05 (default), using backtrack and a dynamic seeding (18). To be able to quantitatively assess connectivity, spherical-deconvolution informed filtering of tractograms (SIFT2) was applied to the resulting tractograms (18). The connectivity matrix was constructed using a robust approach, a 2 mm radial search at the end of the streamline was performed to allow the tracts to reach the grey matter parcellation (19). The final connectivity matrices were multiplied by the *m* coefficient obtained during the SIFT2 process. These connectomes gave a quantification of the SIFT2 weights (referred to as the streamline counts), and the mean FA of connections, between both the left and right amygdala to 41 unilateral regions of interest defined through M-CRIB parcellation.

##### Covariate selection and inclusion

Based on literature, we considered the following potential confounders given their relationships with neonatal brain structure and/or neurodevelopment: GA at birth (20,21), birthweight and birthweight z-score (22), sex (22–25), breast milk feeding (26,27), infant ethnicity (28), maternal age (29), maternal BMI at pregnancy booking (30,31), maternal smoking during pregnancy (32), maternal postnatal depression (33), and maternal highest educational qualification (34,35). Coding and type of these variables are detailed in Supplementary Table 1. We used Pearson correlation to investigate correlations between continuous variables and neurodevelopmental outcomes, and two-sample t-tests to compare outcomes between groups defined by categorical nominal variables. Variables that were nominally significantly ($p < 0.05$) associated with at least one outcome measure were controlled for in downstream statistical analyses. These were GA at birth, sex, ethnicity, maternal postnatal depression score (dichotomised as ≤10), maternal age and maternal education. We additionally adjusted for GA at scan due to its strong correlation with brain MRI metrics.

##### Maternal hair cortisol concentration

Sampling and measurement of maternal hair cortisol concentration is detailed in our previous publication (14). Briefly, maternal hair was sampled within 10 days of delivery. Hair was cut close to the scalp, at the posterior vertex, and stored in aluminium foil at −20°C. The proximal 3 cm of hair were analysed by liquid chromatography-tandem mass spectrometry (LC-MS/MS), at Dresden Lab Service GmbH (Dresden, Germany), using an established protocol (36).

#### SUPPLEMENTARY TABLES

Please note that Tables S2 and S3 are provided in a separate Excel document.

***Table S1. Coding and type of potential covariates collected through questionnaires and medical records.***

| Variable | Coded as | Type |
|---|---|---|
| Infant sex | Male or female | Categorical nominal |
| Infant gestational age at birth | Weeks | Continuous |
| Infant birthweight | Grams | Continuous |
| Birthweight z-score | Weight z-score calculated based on the International Fetal and Newborn Growth Consortium for the 21st Century (INTERGROWTH-21st) standards for preterm infants (37) | Continuous |
| Infant gestational age at MRI scan | Weeks | Continuous |
| Infant ethnicity | Any white background or any other ethnic group | Categorical nominal |
| Infant feeding at discharge | Exclusive breastmilk/mixed feeding or exclusive formula feeding | Categorical nominal |
| Maternal age | Years | Continuous |
| Maternal BMI at pregnancy booking | kg/m2 | Continuous |
| Maternal education (i.e. mother's final educational qualification) | Data was obtained as following:<br>• 1 = none<br>• 2 = 1-4 National 5s / Standard Grades / General Certificate of Secondary Education<br>• 3 = > 5 National 5s / Standard Grades / General Certificate of Secondary Education<br>• 4 = A levels / Highers / equivalent<br>• 5 = College qualification (e.g. National Certificate, Higher National Certificate, Higher National Diploma)<br>• 6 = University undergraduate degree<br>• 7 = University postgraduate degree<br>From this data we created a dichotomous variable by combining brackets 1-5 and 6-7 to indicate whether the mother had obtained a university/postgraduate degree. | Categorical nominal |
| Maternal risk for postnatal depression | Score of 10 or higher on the self-reported Edinburgh Postnatal Depression Scale (38) at the MRI appointment at term-equivalent age. | Categorical nominal |
| Maternal smoking | Current smoker or never/ex-smoker | Categorical nominal |

### SUPPLEMENTARY FIGURES

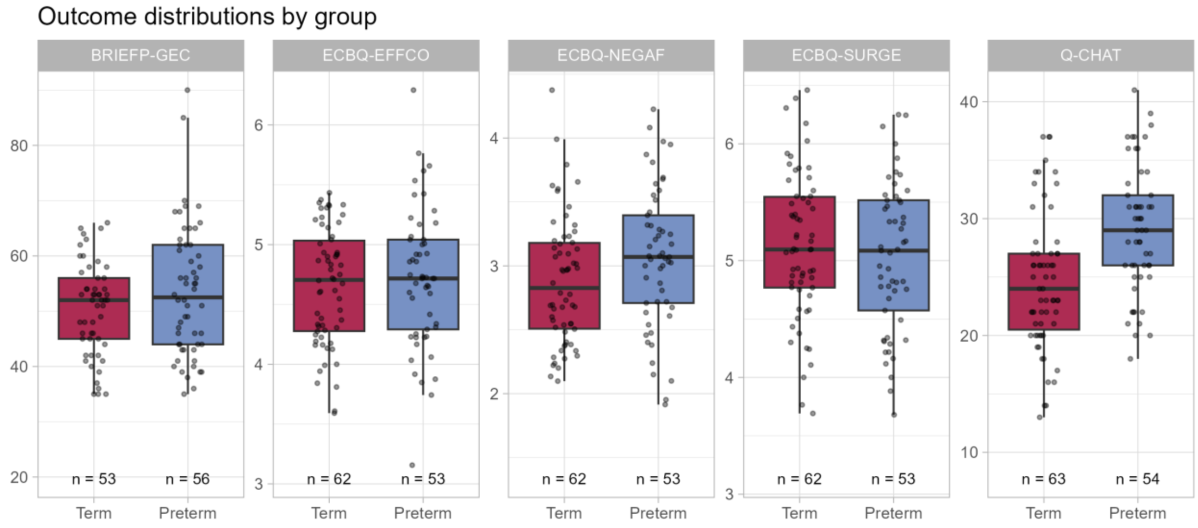


***Figure S1. Distribution of 2-year outcomes in term and preterm groups.*** *Welch t-test indicated a higher Q-CHAT score ($t = -4.345$, $p = 2.99 \times 10^{-5}$) and a slightly higher ECBQ negative affectivity trait in preterm compared to the term group ($t = -1.901$, $p = 0.060$). BRIEF-P = Behavior Rating Inventory of Executive Function, Preschool, GEC = global executive composite, Q-CHAT = Quantitative Checklist for Autism in Toddlers; ECBQ = Early Childhood Behavior Questionnaire; EFFCO = effortful control, NEGAF = negative affectivity, SURGE = surgency.*

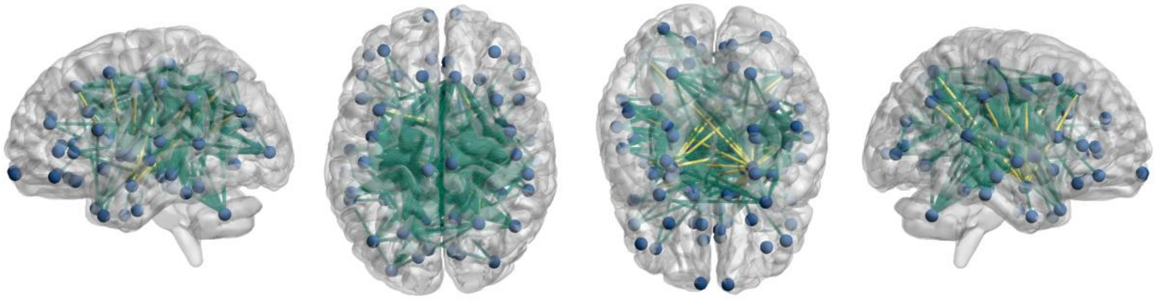

***Figure S2. Illustration of network-based statistics results for ECBQ surgency dimension in preterm infants.*** *Connections that appear as significant in at least 33.3% of experiments (mostly at test-statistic thresholds from 1.5 to 2.2) are shown; amygdala connections are shown in yellow. The connections are between left amygdala and left paracentral gyrus, right inferior parietal cortex, right isthmus cingulate, right postcentral gyrus and right precuneus, and between right amygdala and left caudal middle frontal cortex, left inferior parietal cortex, left precentral gyrus, left precuneus, left superior frontal cortex, left superior parietal cortex, left supramarginal gyrus, right caudate, right paracentral gyrus, right postcentral gyrus, right precentral gyrus, right superior frontal cortex, right superior parietal cortex, and right supramarginal gyrus.*

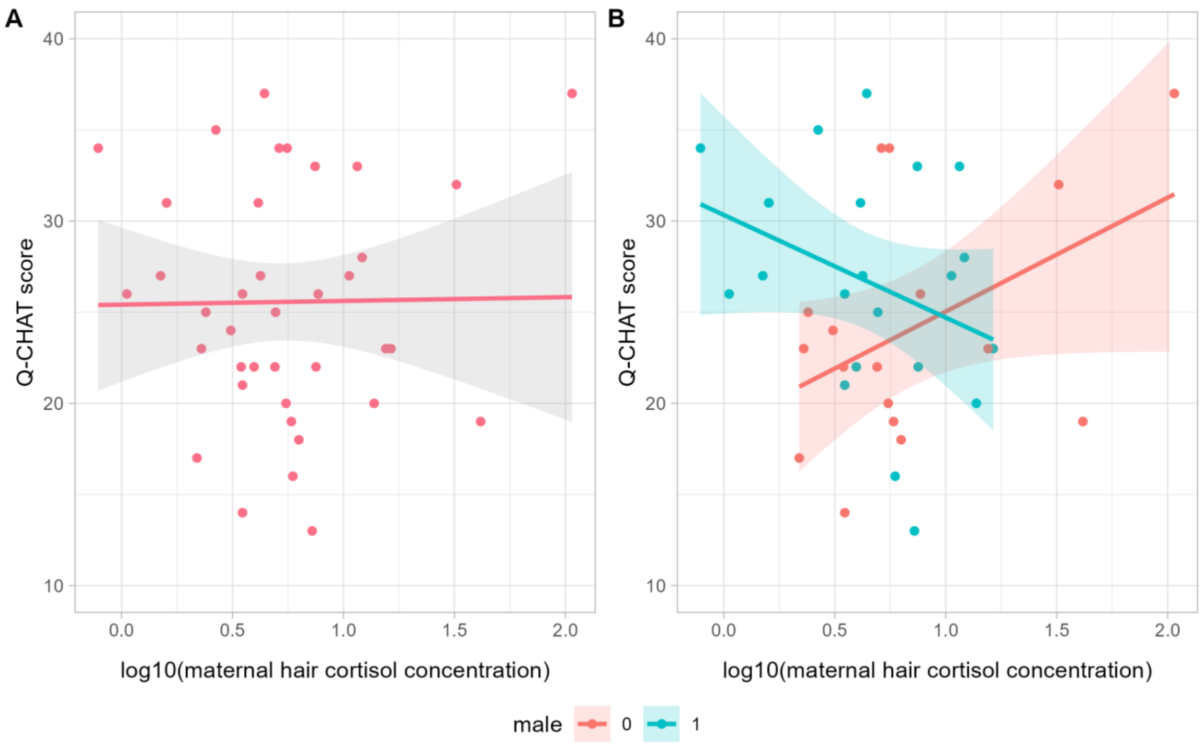


***Figure S3. Maternal hair cortisol and Q-CHAT.*** *(A) There is no correlation between Q-CHAT score and maternal hair cortisol concentration across the whole subsample (n=39). (B) Sex-specific correlations between maternal hair cortisol concentration and Q-CHAT ($\beta_{interaction}$ =-0.661, p=0.043, adjusted for GA at birth, n=39).*
